# The COVID-19 Community Research Partnership: Objectives, Study Design, Baseline Recruitment, and Retention

**DOI:** 10.1101/2022.02.09.22270272

**Authors:** The COVID-19 Community Research Partnership, John Sanders

**Affiliations:** Wake Forest School of Medicine; Wake Forest School of Medicine, Winston Salem, North Carolina, USA

## Abstract

The COVID-19 Community Research Partnership (CCRP) is a multisite surveillance platform designed to characterize the epidemiology of the SARS-CoV-2 pandemic. This manuscript describes the CCRP study design and methodology. The CCRP includes two prospective cohorts, one with six health systems in the mid-Atlantic and southern United States, and the other with six health systems in North Carolina. With enrollment beginning April 2020, sites invited persons within their healthcare systems as well as community members to participate in daily surveillance for symptoms of COVID-like illnesses, testing and risk behaviors. Participants with electronic health records were also asked to volunteer data access. Subsets of participants, representative of the general population and including oversampling of populations of interest, were selected for repeated at home serology testing. By October 2021, 65,739 participants (62,261 adult and 3,478 pediatric) were enrolled with 89% providing syndromic data, 74% providing EHR data, and 70% participating in one of two serology sub-studies. An average of 62% of participants completed a daily survey at least once a week, and 55% of serology kits were returned. The CCRP provides rich regional epidemiologic data and opportunities to more fully characterize the risks and sequelae of SARS-CoV-2 infection.

## INTRODUCTION

The emergence of the SARS-CoV-2 pandemic led to the urgent need for evidence-based answers to pressing clinical and public health questions. Initial efforts focused on counting and describing severe cases or monitoring clinical volumes and establishing patient registries;(1) however, these methods are limited.(1-4) While essential for public health planning, these study designs are insufficient to answer the following essential questions:

1. What is the spectrum of SARS-CoV-2 disease in the population? The importance of asymptomatic carriers and mild cases on community spread cannot be overstated, and assessing only those needing hospital care or those presenting for testing cannot address this issue.
2. Who is at risk from SARS-CoV-2 transmission and disease? Larger studies of more diverse populations followed over time are needed to investigate disparities by race/ethnicity, healthcare worker status, age, and comorbidities.
3. What are the correlates of protection against SARS-CoV-2 infection and outcomes? Uncertainty remains about the level of protection from natural infection with varying degrees of symptom severity, and the real-world efficacy of the various vaccines, especially as new variants emerge and become prevalent.
4. What are the sequelae of SARS-CoV-2 infection at the population level? By focusing entirely on hospitalization and death, many of the impacts of SARS-CoV-2 infection in the community may be missed.

Real-world cohorts were urgently needed to address these growing research gaps. The COVID-19 Community Research Partnership (CCRP) was formed to address these limitations. The CCRP is a health systems-based surveillance platform that includes a complementary triad of regular electronic surveys, longitudinal serologic testing, and Electronic Health Record (EHR) data designed to more accurately characterize the epidemiology of the SARS-CoV-2 pandemic. The CCRP leverages tools available through mobile technologies and at-home laboratory testing across large sample sizes at multiple sites(1) to characterize the epidemiologic cohorts. In this manuscript, we describe the study design and methodology of the CCRP.

## STUDY OBJECTIVES

The primary objective of the CCRP was to collect more comprehensive data on the SARS-CoV-2 pandemic in the US using a cohort approach in order to provide timely and robust evidence for clinical and public health decision making. Specifically, the goal of the CCRP was to collect longitudinal information from four data sources: 1) daily self-reporting of symptoms, testing, and behaviors, 2) comorbidities, medications, medical encounters, and hospitalizations documented in the electronic health record (EHR), 3) SARS-CoV-2 serology and viral surveillance results from targeted subsets within the study, and 4) intermittent participant questionnaires designed to capture supplementary data. The combination of these data elements provided an opportunity to address a diverse set of questions about the SARS-CoV-2 pandemic across the epidemic curve, including the full spectrum of disease, assessment of risk for infection and clinical disease in various subgroups, assessment of population correlates of protection, and identification of potential sequelae.

## METHODS

### Overall Cohort Structure

The COVID-19 Community Research Partnership is composed of two prospective cohorts one located in North Carolina (NC State Coalition) funded by the State of North Carolina through the CARES Act, and the other in the mid-Atlantic and southern United States (National Coalition) funded by the Centers for Disease Control and Prevention (CDC). Each cohort includes six sites, with two sites included in both cohorts.

### Study Sites

The sites in the National Coalition of the CCRP include Wake Forest Baptist Health (WFBH), Atrium Health, MedStar Health/Georgetown University, the University of Maryland School of Medicine/University of Maryland Medical System, the University of Mississippi Medical Center, and Tulane University. The sites in the NC State Coalition of the CCRP include WFBH, Atrium Health WakeMed Health, New Hanover Regional Medical Center, Vidant Health, and Campbell University. WFBH and Atrium Health in both cohorts (Figure 1). Supplemental Table 1 provides specifics of site eligibility, start dates, and recruitment where different from that of the overall CCRP. The two cohorts are linked by common design features and data coordination but differ in serologic testing platforms and sub-study activity (described in detail below). All sites utilize a common Data Coordinating Center (DCC) at the George Washington University Biostatistics Center, a common research program management office at Vysnova Partners, Inc., and Call Centers that respond to participant inquiries and technical problems. Further details on study organization are included in Supplemental Figure 1. Data from the two cohorts are merged in a common database, with tags indicating which participants from WFBH and Atrium are included in each cohort, allowing analyses within the entire cohort or within each cohort separately as required.

**Figure 1:**
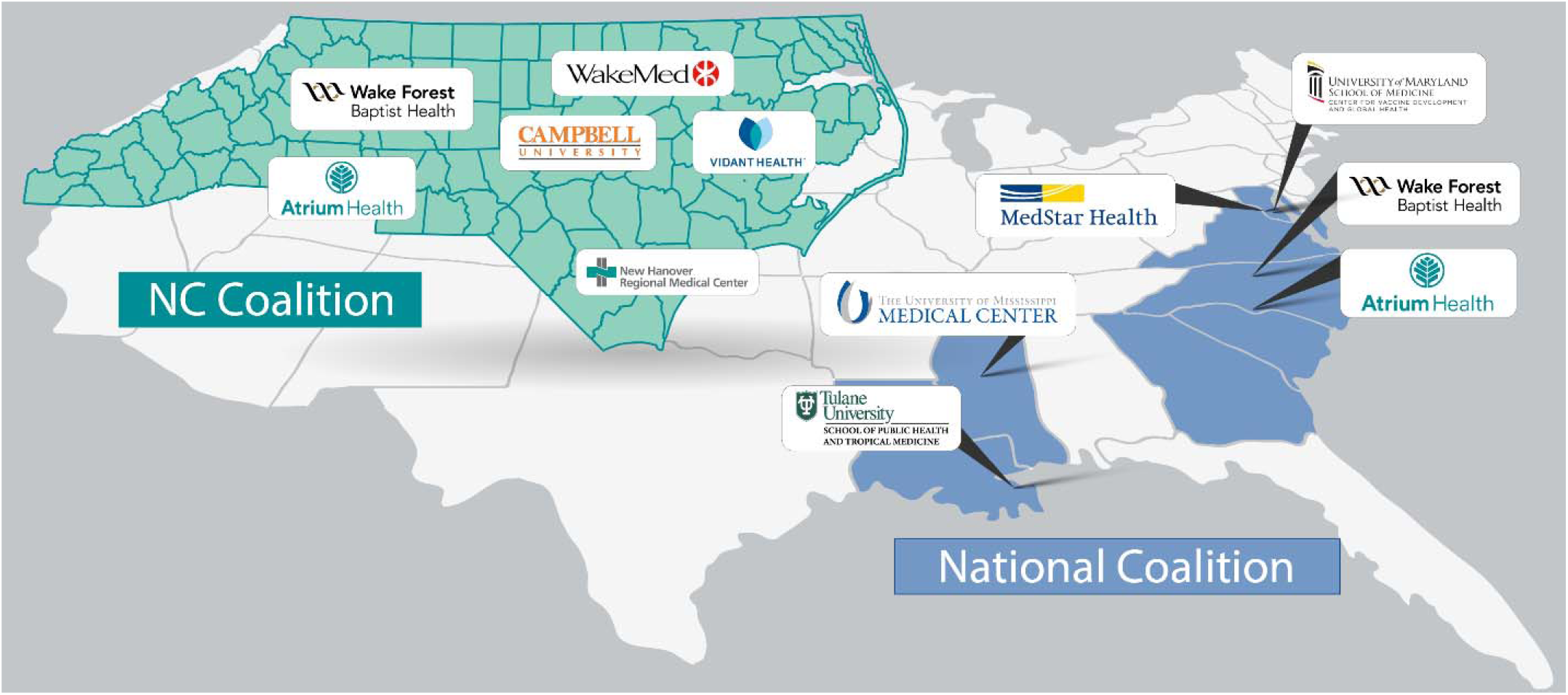
COVID-19 Community Research Partnership Site Locations

### Recruitment

Eligibility for the CCRP study was broad and included most adults, with the only inclusion requirements being an active email address and cell phone with data capabilities. In addition to the adults recruited to participate in the two cohorts, pediatric participants were recruited to the NC Coalition. Initially, invitations to participate in the CCRP were sent out through participating healthcare systems beginning in March 2020 at WFBH and progressively including additional sites as funding allowed. Healthcare workers were oversampled. Additionally, sites had the option to invite community members outside of the participating healthcare system to participate in the CCRP. Recruitment included multiple methods: 1) email or texts using the health systems’ patient communication portals (e.g., MyChart (5); 2) social media and public relations campaigns led by individual sites; 3) virtual and in-person community outreach; and 4) a website accessible to the public with information about the CCRP (http://www.covid19communitystudy.org/index.html). See Supplemental Table 1 for additional detail about recruitment strategy by site.

### Daily Syndromic Surveillance and Supplemental Surveys

Following informed consent provided through secure web-based systems, participants completed a onetime enrollment questionnaire (Supplemental Figure 2) which was immediately followed by a survey questionnaire which collected basic demographics (age, sex, and race/ethnicity), address, and if they were a current healthcare worker (Supplemental Figure 3). Following the consent and enrollment process, participants completed daily electronic surveys via computer or smartphone using one of two electronic systems, the Oracle Patient Monitoring System (PMS) (Oracle Corporation, Redwood Shores, California) or the SneezSafe application (Sneez, LLC, Winston Salem, NC, USA).

The daily syndromic survey asked participants to log the presence or absence of a variety of symptoms related to Covid-19 infection (Supplemental Figure 4). Additional questions included mask use, care-seeking behaviors, known COVID-19 exposures, and self-reported SARS-CoV-2 testing results. Healthcare workers reporting COVID-19 contacts also provide data on frequency and type of personal protective equipment (PPE) use at the time of the potential exposure. A question about COVID-19 vaccination status and vaccine type was added in September 2020. Daily data entry typically took <30 seconds.

In addition, all CCRP participants completed periodic supplemental surveys through Oracle PMS or SneezSafe. Surveys were developed to be responsive to changing trends and newly arising research questions during the pandemic. To date, supplemental surveys have focused on risk mitigation behavior such as mask use during the Thanksgiving and winter holidays(6); intentions and hesitancy to vaccinate; a household survey concerning family members (e.g., number, medical history, family income); and an individual adult survey exploring comorbidities, health behaviors (e.g., flu vaccination, alcohol, smoking), education, and occupation. These surveys will be described in detail in relevant manuscripts.

### Electronic Health Record

A subset of CCRP participants, 67%, agreed to have their personal EHR data extracted and included in the study. Participants were matched to their EHR by each health system using patient-provided characteristics including name, birth date, and address, and then authenticated. To capture health conditions prevalent before the pandemic and ensure that pertinent diagnoses were not missed due to potentially reduced health care use during the pandemic, the EHR was abstracted from a look-back period that started in January 2018. Key EHR data are extracted at quarterly intervals to identify patient characteristics, health service utilization, diagnoses, medications, procedures, and lab test results throughout the study. The majority of EHR content is made up of readily identifiable structured data fields, extracted using formats based on the Fast Healthcare Interoperability Resources (FHIR) standard, and use established standards (e.g. ICD10 diagnosis codes, procedure codes, medications, patient demographics, insurance, vital signs, test results, health behaviors) where possible. Data are checked for quality to remove impossible values, and harmonized across participating healthcare systems to the extent possible to enable meaningful analysis over the CCRP study population. Health conditions of interest extracted from the EHR are listed in Supplemental Table 2.

### Longitudinal Serology Testing

A subset of the participants from the CCRP were selected for repeated, longitudinal, at-home serologic sampling. Selected participants were eligible to receive at least six tests over the 12-month period. The selection strategy for testing and the test platform utilized differed between the National Coalition and the NC State Coalition as further described.

### National Coalition Serology Testing

Within the National Coalition, investigators chose serology participants from the counties in which the participating healthcare systems provides service. The counties in the catchment area of Atrium Health and WFBH were grouped into a Southeastern area, those of Mississippi University and Tulane University into a Deep South area, and those of University of Maryland and MedStar Health/Georgetown University into a Mid-Atlantic area. The counties in those catchment areas were characterized as urban, suburban, or rural. Participants were selected for serologic testing to represent the overall demographics (sex, race/ethnicity, age) of the area based on the 2017 American Community Survey (ACS) census.(7) A target bin size was determined based on the demographics of the area, and enrollment in a stratum was capped when the target bin size was reached. Additional participants were selected by oversampling subgroups considered to be at high risk for infection or complications from infection (predominantly healthcare workers and minority populations), and a small number of additional participants were selected to allow for flexibility throughout the enrollment process.

Serologic testing for the National Coalition utilized at-home collection of dried blood spots on a Whatman 5-spot DBS specimen card. LabCorp delivered a study-branded kit with instructions for collection, two lancets, and a self-addressed return envelope to participants. Once returned, LabCorp performed EUROIMMUN enzyme-linked immunosorbent assay (ELISA) from the dried blood spot targeting the SARS-CoV-2 spike protein. This test has been granted Emergency Use Authorization (EUA) from the FDA for testing on venous blood(8) and was internally validated for use with dried blood spot specimens. Testing performance is described in Supplemental Table 3. This test was expected to be positive following natural infection or vaccination. For further discrimination between natural or vaccine-induced antibodies, positive tests on the EUROIMMUN assay automatically reflexed to testing for nucleocapsid antibodies utilizing the EUA-approved Roche Elecsys Anti-SARS-CoV-2 ECLIA. A natural infection would be Roche assay positive and vaccine-induced response Roche assay negative. The dried blood spot cards with unused spots have been stored in a biorepository for future ancillary studies.

### NC State Coalition Serology Testing

NC State Coalition serologic testing began in April 2020, before the formation of the National Coalition. The initial cadence of testing for the NC State Coalition was every other month, with additional testing added depending on available funding. Initially, participants were mailed a simple micro-sampling devise to collect a total of 40microL blood using two volumetric absorptive swab tips (Neoteryx LLC, Torrance, California, USA). The tips were returned by pre-addressed mail to our laboratory for further processing using a one of two lateral flow assays (LFA) detecting IgM and/or IgG antibodies to SARS-CoV-2 (Syntron, Syntron Bio Research Inc., Carlsbad, California, USA or Innovita, Beijing Innovita Biological Technology, China). Later, we switched to at-home serologic testing for the NC State Coalition using a lateral flow assay (LFA), the Scanwell SARS-CoV-2 IgM IgG Test (Teco Diagnostics). Scanwell Health shipped the LFAs by mail to be performed at home by participants, and the result was recorded and interpreted via an FDA-approved smartphone application developed by Scanwell Health. Interpretation by Scanwell staff involved reading a standard 3-line result for IgM, IgG and control. Test performance (9) is describe in Supplemental Table 3.

### NC State Coalition Virology Testing

A subset of adult and pediatric participants in the NC State Coalition who consented to at-home testing completed at-home PCR-based viral surveillance. Selected adult and pediatric participants received four viral test kits and were asked to collect a sample weekly for four weeks. If the participant experienced persistent or worsening cough, fever, or loss of taste, they were asked to complete and return the test kit before the end of the week.

Study-branded mailers and instructional videos were developed to enable participants to conduct their own sampling at home. For each test, the participant or their caregiver collected an oral specimen of saliva by swabbing (Curative Inc., #K00023) the inside of the mouth until the swab was saturated with saliva. The swab was then placed into a capped tube (Curative Inc., #K00016) containing DNA/RNA Shield (Zymo Research, #R1100) that in turn was placed in a return mailer. The lysis buffer inactivates the SARS-CoV-2 virus immediately reducing biosafety and shipping concerns. The molecular detection assay uses RT-PCR based on the CDC EUA adapted by Curative-Korva.(10) As the regulatory environment was in a fluid state, the latest guidance on available EUA assays for SARS-CoV-2 was generally followed.

### Study Participation and Retention

Although participants could join at any time, adult recruitment ended in April 2021 and pediatric recruitment ended in September 2021. By October 2021, 65,739 participants (62,261 adults and 3,478 pediatric) were enrolled in the CCRP, with 89% providing syndromic data, 74% providing EHR data, and 70% participating in one of the two serology sub-studies. Accumulated enrollment in the CCRP and serology substudies by demographic characteristics is shown in Table 1, and demographic distribution by study site is shown in Supplemental Table 4. By October 2021, an average of 62% of participants complete a daily survey at least once a week, and 55% of serology kits had been completed and returned. Retention by cohort for daily collection of syndromic data is shown in Figure 2.

**Table 1:** Enrollment Characteristics in the COVID-19 Community Research Partnership and Serology Substudies

|  | **Number (Percent) of**<br>**Enrolled** | **Number (Percent)**<br>**Enrolled in CDC**<br>**Serologic Substudy** | **Number (Percent)**<br>**Enrolled in NC State**<br>**Serology Substudy** |
| --- | --- | --- | --- |
| **Total** | 62261 | 24965 | 21849 |
| **Age** |  |  |  |
| 18-29 | 6427 (10.3%) | 1829 (7.3%) | 1872 (8.6%) |
| 30-39 | 11180 (18.0%) | 4507 (18.1%) | 4058 (18.6%) |
| 40-49 | 11806 (19.0%) | 4275 (17.1%) | 4796 (22.0%) |
| 50-59 | 12578 (20.2%) | 4363 (17.5%) | 4991 (22.8%) |
| 60-69 | 12689 (20.4%) | 5621 (22.5%) | 4456 (20.4%) |
| 70-79 | 6546 (10.5%) | 3775 (15.1%) | 1476 (6.8%) |
| >=80 | 990 (1.6%) | 595 (2.4%) | 200 (0.9%) |
| **Sex** |  |  |  |
| Female | 40915 (65.8%) | 16121 (64.6%) | 15974 (73.1%) |
| Male | 18470 (29.7%) | 8844 (35.4%) | 5728 (26.2%) |
| Not Declared | 2831 (4.6%) | 0 (0.0%) | 147 (0.7%) |
| **Race/Ethnicity** |  |  |  |
| White (not<br>Hispanic/Latino) | 50485 (81.1%) | 20203 (80.9%) | 20346 (93.1%) |
| Black or African<br>American | 4102 (6.6%) | 2235 (9.0%) | 550 (2.5%) |
| American Indian or<br>Alaskan | 221 (0.4%) | 66 (0.3%) | 86 (0.4%) |
| Asian or Pacific Islander | 1329 (2.1%) | 840 (3.4%) | 194 (0.9%) |
| Hispanic or Latino | 1602 (2.6%) | 982 (3.9%) | 231 (1.1%) |
| Mixed Ethnicity | 918 (1.5%) | 574 (2.3%) | 163 (0.7%) |
| Not Declared/Specified | 3559 (5.7%) | 65 (0.3%) | 279 (1.3%) |
| **Geography/Site** |  |  |  |
| Wake Forest Baptist<br>Health | 26131 (42.0%) | 8017 (32.1%) | 12220 (55.9%) |
| University of Maryland | 5905 (9.5%) | 3840 (15.4%) | 0 (0.0%) |
| Atrium | 9557 (15.4%) | 3922 (15.7%) | 4099 (18.8%) |
| MedStar Health | 12995 (20.9%) | 8779 (35.2%) | 0 (0.0%) |
| Tulane | 244 (0.4%) | 187 (0.7%) | 0 (0.0%) |
| University of Mississippi | 478 (0.8%) | 220 (0.9%) | 0 (0.0%) |
| Campbell | 889 (1.4%) | 0 (0.0%) | 536 (2.5%) |
| New Hanover | 810 (1.3%) | 0 (0.0%) | 761 (3.5%) |
| Vidant Health | 1671 (2.7%) | 0 (0.0%) | 990 (4.5%) |
| Wake Med | 3536 (5.7%) | 0 (0.0%) | 3243 (14.8%) |

**Figure 2:**
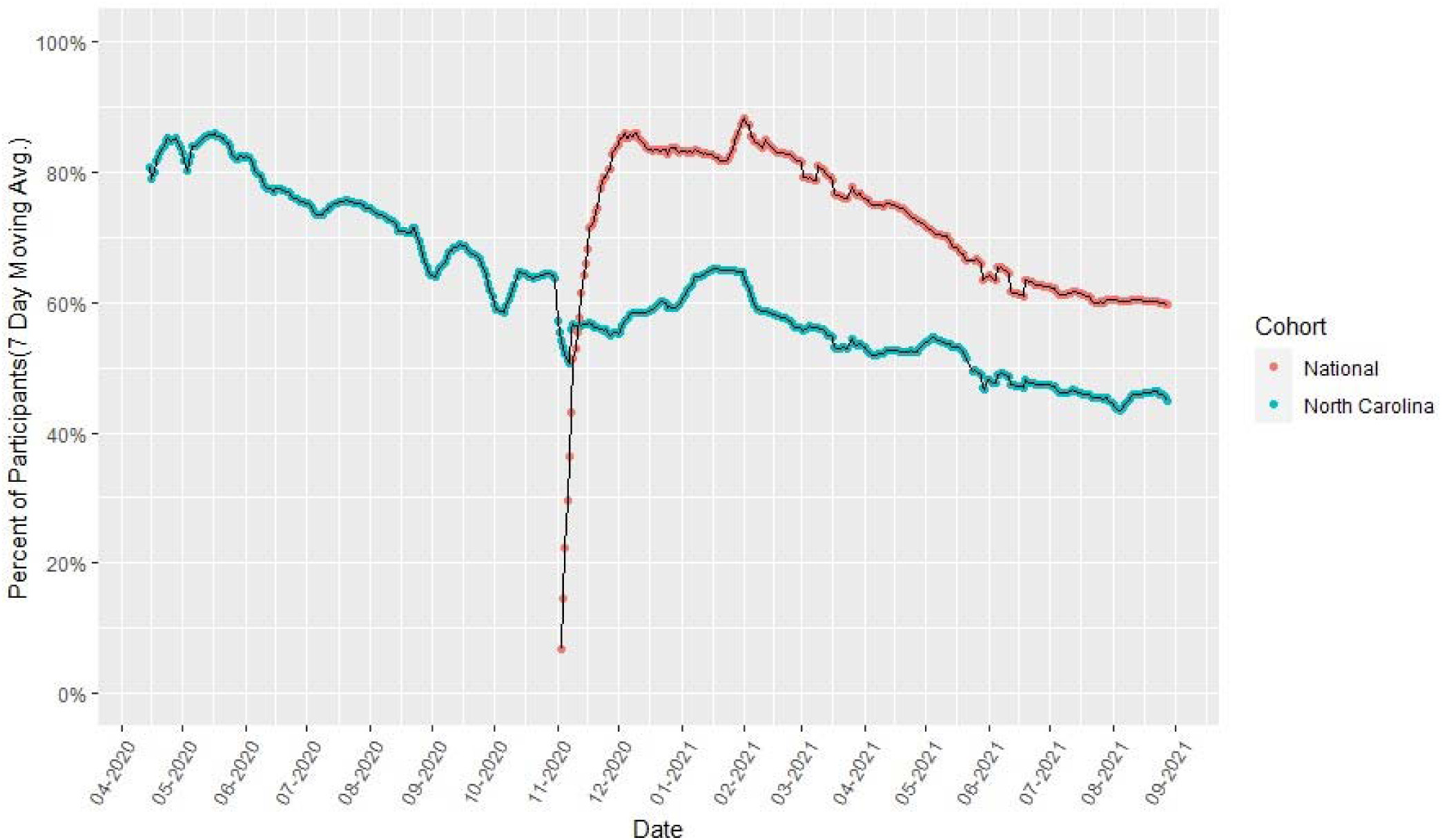
Participant Retention in COVID-like Illness Surveillance by Cohort and Study Date Data points represent the percentage of participants in the National and NC State Cohort completing daily symptom logs by calendar date smoothed using a 7-day moving average and removing data on 26 and 27 April 2020, 24 and 25 March 2021, and 22 to 23 May 2021 due to computer downtime. For NC State cohort, the starting retention rate was about 80% calculated from the first date with >= 200 enrollees (15 April 2020). By 28 August 2021, the rate was 45%. For the National cohort, the starting retention rate was about 80% based on the first date where we had at least 200 enrollees (26 November 2020). By 28 August 2021, the rate was 60%.

### Ethical Considerations

The CCRP is conducted in accordance with all US federal regulations regarding protection of human subjects. The Institutional Review Board (IRB) at Wake Forest Baptist Health serves as the central or reliance IRB. Every site IRB and the DCC have an Authorization Agreement with WFBH IRB. Any changes to protocol or implementation are notified to the site IRB for processing as needed.

A limited waiver of Health Insurance Portability and Accountability Act (HIPAA) authorization was granted by the IRB to identify potential subjects for recruitment, as allowed under 45 CFR 164.512. All data are kept confidential to the extent permitted by applicable state, local and federal laws, and the data are stored in secured HIPAA-compliant electronic databases at George Washington University, Scanwell Health, and Oracle with password-protected access. All data are stored on secure servers in the United States. To ensure confidentiality, all data for analysis are secured, and a coded number is used to identify participants.

Participants were provided an opportunity to consent to different aspects of the study separately, including access to their EHR, participation in symptom surveillance, serosurveillance and viral testing, and storage of residual samples. Based on EHR access, the consent was designed to serve as HIPAA authorization.

Among eligible persons or their guardians, a secure link was provided to online informed consent and enrollment through one of three electronic platforms. Sites had the option to host their own system using REDCap (Research Electronic Data Capture), a software toolset and workflow methodology for electronic collection and management of data (8,9). An alternative consent and enrollment process used by several sites was provided by the DCC using a cybersecure, internally-developed data entry and management system known as MIDAS (Multimodal Integrated Data Acquisition System). A third option was provided through the SneezSafe platform, a cybersecure, phone, tablet or computer -based application consent and symptom-screening tool offered by Sneez, LLC. Information describing the study, common consent language, and enrollment steps were conserved regardless of platform used. All PII was stored separately from research data, and accessible only to individuals requiring access (e.g., those responsible for mailing serology or virology test kits).

At enrollment, during informed consent, and upon receipt of a test result, participants were informed that the serology and viral diagnostic tests are for research purposes only, and should not be interpreted or used for clinical diagnostic purposes. For each serology and virology result, a standardized, secure, IRB-approved message on the interpretation of a positive, negative, or equivocal result is sent to the participant, which also includes the contact information for the Call Center. Concerned volunteers were encouraged to contact the Call Center for test result information and counseling by a trained clinical professional (e.g., nurse) or, if required due to complex medical concerns, a study physician.

### Data Management

A database of all study data is maintained and managed at the DCC. Data from the daily syndromic reporting and periodic supplemental questionnaires collected through SneezSafe, and all serology and virology test results, are securely loaded into the Oracle PMS. All data are downloaded three times weekly to the DCC’s secure server. EHR data are provided approximately every 3 months from each healthcare system directly to the DCC via secure file transfer protocol.

### Data Sharing and Access

An internal, password-protected study website includes dynamic reports for study investigators to track recruitment, retention, and interim results. An external study website provides static reports for study participants and the public to review study demographics and results. De-identified and well-documented databases for the National Coalition are developed by the DCC and provided to the CDC quarterly. At the close of the study, the complete de-identified data will be made publicly available according to CDC policies.

The CCRP Publications and Presentations Committee (P&P) coordinates and reviews all aspects of manuscripts and presentations generated from the CCRP to prevent redundant effort, maintain consistency in reporting, and ensure scientific rigor. The P&P uses a standardized protocol for reviewing and approving manuscript proposals, conference abstracts, presentations, and penultimate drafts of manuscripts before submission to external sources. All data collected as part of the National Coalition also undergoes CDC clearance.

## DISCUSSION

As a large-scale health systems-based cohort combining daily symptom and risk behavior reporting, longitudinal serologic sampling, supplemental surveys, vaccination records, and EHR capture, the COVID-19 Community Research Partnership offers tremendous opportunities to obtain synergies by combining clinically relevant illness, health, and socio-behavioral data. The CCRP design is participant-friendly and embraces modern technologies and practical methods to overcome limitations of more traditional studies. For these reasons, the CCRP provides a unique resource for generating evidence-based answers to questions about the SARS-CoV-2 pandemic in the US.

The CCRP has several limitations. First, the CCRP is not a population-based study and does not consist of a random sample from the populations in the health system catchment areas, so selection bias is likely. In part, this bias is due to the study requirement of having an active email address and certain recruitment methods that relied on access to mobile technology and web or cell connectivity. CCRP participants were overrepresented by older, female, and Non-Hispanic White groups, although serology cohorts were sampled to more closely represent the demographics of surrounding counties. The decision to participate and loss-to-follow up may also have varied by demographics. As such, results may not be generalizable to broader geographic areas or to populations with less access to healthcare or electronic communications. Second, the logistics of providing serology tests to participants in their homes proved challenging, and the selection strategy for testing and the test platform utilized differed between the National Coalition and the NC State Coalition. There were clear tradeoffs between selectively reduced participation from requiring participants to have a smartphone to report the test results from one platform compared to the burden of having to take five bloodspots and return the other test platform. Test cadence was more variable than ideal due to issues with slower test cadence in the NC State Coalition early in the study and the impact of shipping delays and slow participant response throughout the study. Nonetheless, retention in the serology cohorts was high, and this study found that at-home serology testing was generally well accepted and feasible. Third, study retention and continued participation may be impacted by known exposure, infection, and vaccination for SARS-CoV-2. Daily symptom reporting and monthly serology testing over many months can be a burden for participants. In particular, missing data may be sporadic and missing not at random. For example, a participant may stop daily symptom reporting if they do not experience any symptoms for long stretches of time, but may return to daily reporting if they begin to experience symptoms. Similarly, participant engagement with serology testing may decrease after an initial positive test or vaccination.

Despite these limitations, the CCRP has several advantages over more traditional surveillance methods. The cohort study design and longitudinal data collection, including serology, allows the CCRP to assess incidence of SARS-CoV-2 over time.(11) The serology approach from the National Coalition using a nucleocapsid confirmatory test further allowed the CCRP to assess the difference between antibody response to vaccination and natural infection. Daily symptom and self-report of SARS-CoV-2 testing with daily reminders, and a process typically taking less than 30 seconds, minimizes the limitations of typical syndromic surveillance which often suffers from misclassification due to recall bias with longer reporting intervals. By linking these data to longitudinal serology, the CCRP can characterize asymptomatic and minimally symptomatic infections, a key advantage of the CCRP design compared to that of traditional studies. Furthermore, linkage to EHR data allows for the characterization of comorbidities, healthcare encounters, clinical outcomes, and potential sequelae of infection.

The CCRP design offers several advantages in recruiting a large and diverse population. The utilization of health care systems as the central recruiting hubs facilitated communication through patient’s trusted health care systems with the potential to inspire confidence and increase enrollment and participation. Recognizing that some vulnerable populations are not well connected to health care systems, the CCRP included monitoring of enrollment, direct recruitment of under-represented minority groups, and over-sampling of subgroups at increased risk for infection, morbidity and mortality (e.g. health-care workers, minority populations, the elderly), to improve broad representation across socio-demographic groups and geographic regions. The entire study has been conducted remotely utilizing electronic informed consent, electronic/mobile technologies, and in-home sample collection and testing which enhances involvement of participants with poor access to the healthcare facilities and is supportive of social distancing measures.

## Conclusions

The CCRP is providing new and timely information about the rapidly changing SARS-CoV-2 pandemic in the US. Symptom surveillance tied to longitudinal serology makes it possible to describe the full spectrum of disease associated with SARS-CoV-2 infection, from asymptomatic or minimally symptomatic cases to those with more severe symptoms. Serology data allows investigators to calculate the seroprevalence and incidence within demographic subgroups, and along with EHR data, to assess risk of SARS-CoV-2 infection and severity associated with pre-existing medical conditions, and the impact of SARS-CoV-2 infection on other outcomes. Finally, the CCRP provides essential evidence of real world vaccine efficacy and changing SARS-CoV-2 mitigation behaviors related to emerging recommendations and events. It is hoped that results from the CCRP will contribute to clinical and public health recommendations and practice to help better understand and control the SARS-CoV-2 pandemic.

## Data Availability

All data produced in the present study are available upon reasonable request to the corresponding author.

## Acknowledgements

The CCRP investigators thank the 64,481 participants who collected data on a daily basis and the 42,708 participants who also collected serologic specimens monthly.

## FIGURES AND TABLES

Supplemental Figure 1: Organizational Structure of the COVID-19 Community Research Partnership

Supplemental Figure 2: Enrollment Questionnaire (example from Wake Forest Baptist Health)

Supplemental Figure 3: Registration and Demographics Form

Supplemental Figure 4: Daily COVID-like Illness Report

Supplemental Table 1: Community Research Partnership Collaborating Health Systems, Partipant Eligibility, and Enrolment Strategy

Supplemental Table 2: List of EHR Extracted Data Supplemental Table 3: Characteristics of Serologic Tests

Supplemental Table 4: Demographic Characteristics by Study Site

## ** The COVID-19 Community Research Partnership

**Authorship for The COVID-19 Research Group** (*Site Principal Investigator)

**Wake Forest School of Medicine:** Thomas F Wierzba PhD, MPH*, John Walton Sanders, MD, MPH, David Herrington, MD, MHS, Mark A. Espeland, PhD, MA, John Williamson, PharmD, Morgana Mongraw-Chaffin, PhD, MPH, Alain Bertoni, MD, MPH, Martha A. Alexander-Miller, PhD, Paola Castri, MD, PhD, Allison Mathews, PhD, MA, Iqra Munawar, MS, Austin Lyles Seals, MS, Brian Ostasiewski, Christine Ann Pittman Ballard, MPH, Metin Gurcan, PhD, MS, Alexander Ivanov, MD, Giselle Melendez Zapata, MD, Marlena Westcott, PhD, Karen Blinson, Laura Blinson, Mark Mistysyn, Donna Davis, Lynda Doomy, Perrin Henderson, MS, Alicia Jessup, Kimberly Lane, Beverly Levine, PhD, Jessica McCanless, MS, Sharon McDaniel, Kathryn Melius, MS, Christine O’Neill, Angelina Pack, RN, Ritu Rathee, RN, Scott Rushing, Jennifer Sheets, Sandra Soots, RN, Michele Wall, Samantha Wheeler, John White, Lisa Wilkerson, Rebekah Wilson, Kenneth Wilson, Deb Burcombe, Georgia Saylor, Megan Lunn, Karina Ordonez, Ashley O’Steen, MS, Leigh Wagner.

**Atrium Health:** Michael S. Runyon MD, MPH*, Lewis H. McCurdy MD*, Michael A. Gibbs, MD, Yhenneko J. Taylor, PhD, Lydia Calamari, MD, Hazel Tapp, PhD, Amina Ahmed, MD, Michael Brennan, DDS, Lindsay Munn, PhD RN, Keerti L. Dantuluri, MD, Timothy Hetherington, MS, Lauren C. Lu, Connell Dunn, Melanie Hogg, MS, CCRA, Andrea Price, Marina Leonidas, Melinda Manning, Whitney Rossman, MS, Frank X. Gohs, MS, Anna Harris, MPH, Jennifer S. Priem, PhD, MA, Pilar Tochiki, Nicole Wellinsky, Crystal Silva, Tom Ludden PhD, Jackeline Hernandez, MD, Kennisha Spencer, Laura McAlister.

**MedStar Health Research Institute:** William Weintraub MD*, Kristen Miller, DrPH, CPPS*, Chris Washington, Allison Moses, Sarahfaye Dolman, Julissa Zelaya-Portillo, John Erkus, Joseph Blumenthal, Ronald E. Romero Barrientos, Sonita Bennett, Shrenik Shah, Shrey Mathur, Christian Boxley, Paul Kolm, PhD, Ella Franklin, Naheed Ahmed, Moira Larsen.

**Tulane**: Richard Oberhelman MD*, Joseph Keating PhD*, Patricia Kissinger, PhD, John Schieffelin, MD, Joshua Yukich, PhD, Andrew Beron, MPH, Johanna Teigen, MPH.

**University of Maryland School of Medicine:** Karen Kotloff MD*, Wilbur H. Chen MD, MS*, DeAnna Friedman-Klabanoff, MD, Andrea A. Berry, MD, Helen Powell, PhD, Lynnee Roane, MS, RN, Reva Datar, MPH, Colleen Reilly.

**University of Mississippi**: Adolfo Correa MD, PhD*, Bhagyashri Navalkele, MD, Yuan-I Min, PhD, Alexandra Castillo, MPH, Lori Ward, PhD, MS, Robert P. Santos, MD, MSCS, Pramod Anugu, Yan Gao, MPH, Jason Green, Ramona Sandlin, RHIA, Donald Moore, MS, Lemichal Drake, Dorothy Horton, RN, Kendra L. Johnson, MPH, Michael Stover.

**Wake Med Health and Hospitals:** William H. Lagarde MD*, LaMonica Daniel, BSCR.

**New Hanover:** Patrick D. Maguire MD*, Charin L. Hanlon, MD, Lynette McFayden, MSN, CCRP, Isaura Rigo, MD, Kelli Hines, BS, Lindsay Smith, BA, Monique Harris, CCRP, Belinda Lissor, AAS, CCRP, Vivian Cook, MA, MPH, Maddy Eversole, BS, Terry Herrin, BS, Dennis Murphy, RN, Lauren Kinney, BS, Polly Diehl, MS, RHIA, Nicholas Abromitis, BS, Tina St. Pierre, BS, Bill Heckman, Denise Evans, Julian March, BA, Ben Whitlock, CPA, MSA, Wendy Moore, BS, AAS, Sarah Arthur, MSW, LCSW, Joseph Conway.

**Vidant Health:** Thomas R. Gallaher MD*, Mathew Johanson, MHA, CHFP, Sawyer Brown, MHA, Tina Dixon, MPA, Martha Reavis, Shakira Henderson, PhD, DNP, MS, MPH, Michael Zimmer, PhD, Danielle Oliver, Kasheta Jackson, DNP, RN, Monica Menon, MHA, Brandon Bishop, MHA, Rachel Roeth, MHA.

**Campbell University School of Osteopathic Medicine**: Robin King-Thiele DO*, Terri S. Hamrick PhD*, Abdalla Ihmeidan, MHA, Amy Hinkelman, PhD, Chika Okafor, MD (Cape Fear Valley Medical Center), Regina B. Bray Brown, MD, Amber Brewster, MD, Danius Bouyi, DO, Katrina Lamont, MD, Kazumi Yoshinaga, DO, (Harnett Health System), Poornima Vinod, MD, A. Suman Peela, MD, Giera Denbel, MD, Jason Lo, MD, Mariam Mayet-Khan, DO, Akash Mittal, DO, Reena Motwani, MD, Mohamed Raafat, MD (Southeastern Health System), Evan Schultz, DO, Aderson Joseph, MD, Aalok Parkeh, DO, Dhara Patel, MD, Babar Afridi, DO (Cumberland County Hospital System, Cape Fear Valley).

**George Washington University Data Coordinating Center:** Diane Uschner PhD*, Sharon L. Edelstein, ScM, Michele Santacatterina, PhD, Greg Strylewicz, PhD, Brian Burke, MS, Mihili Gunaratne, MPH, Meghan Turney, MA, Shirley Qin Zhou, MS, Ashley H Tjaden, MPH, Lida Fette, MS, Asare Buahin, Matthew Bott, Sophia Graziani, Ashvi Soni, MS, Guoqing Diao, PhD, Jone Renteria, MS.

**George Washington University Mores Lab:** Christopher Mores, PhD, Abigail Porzucek, MS.

**Oracle Corporation:** Rebecca Laborde, Pranav Acharya.

**Sneez LLC**: Lucy Guill, MBA, Danielle Lamphier, MBA, Anna Schaefer, MSM, William M. Satterwhite, JD, MD.

**Vysnova Partners:** Anne McKeague, PhD, Johnathan Ward, MS, Diana P. Naranjo, MA, Nana Darko, MPH, Kimberly Castellon, BS, Ryan Brink, MSCM, Haris Shehzad, MS, Derek Kuprianov, Douglas McGlasson, MBA, Devin Hayes, BS, Sierra Edwards, MS, Stephane Daphnis, MBA, Britnee Todd, BS.

**Javara Inc:** Atira Goodwin.

**External Advisory Council:** Ruth Berkelman, MD, Emory, Kimberly Hanson, MD, U of Utah, Scott Zeger, PhD, Johns Hopkins, Cavan Reilly, PhD, U. of Minnesota, Kathy Edwards, MD, Vanderbilt, Helene Gayle, MD MPH, Chicago Community Trust, Stephen Redd.

