## Supplementary figures and images for "The COVID-19 Community Research Partnership: Objectives, Study Design, Baseline Recruitment, and Retention"

### Supplemental Figure 1

Supplemental Figure 1. Organizational Structure of the COVID-19 Community Research Partnership

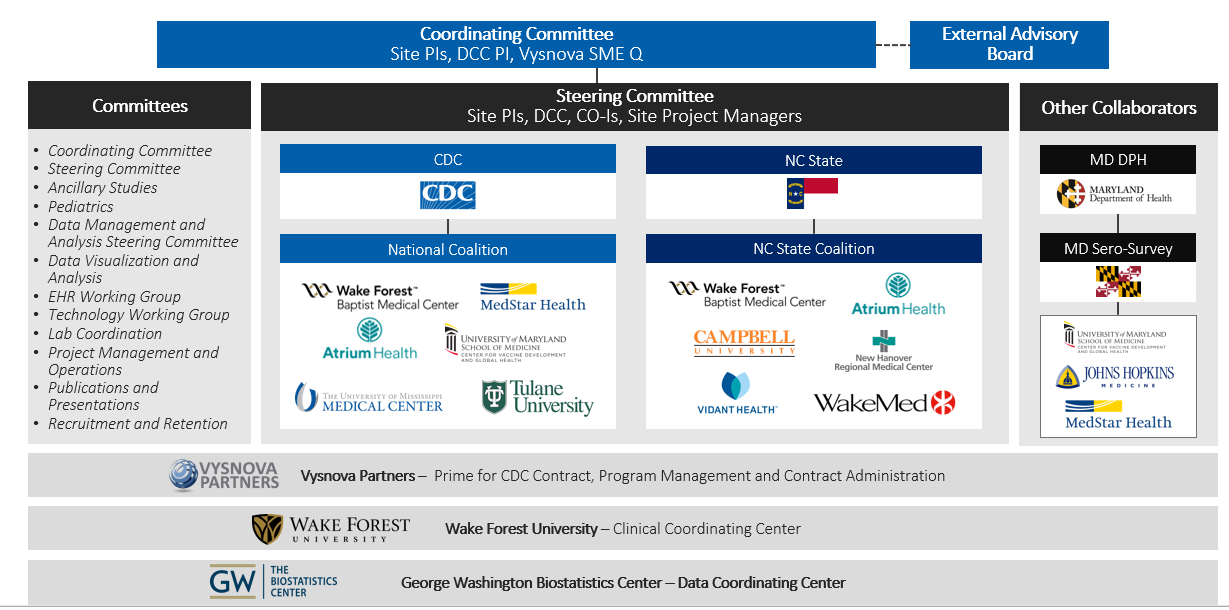

### Supplemental Figure 2

Supplemental Figure 2: Enrollment Questionnaire (example from Wake Forest Baptist Health)

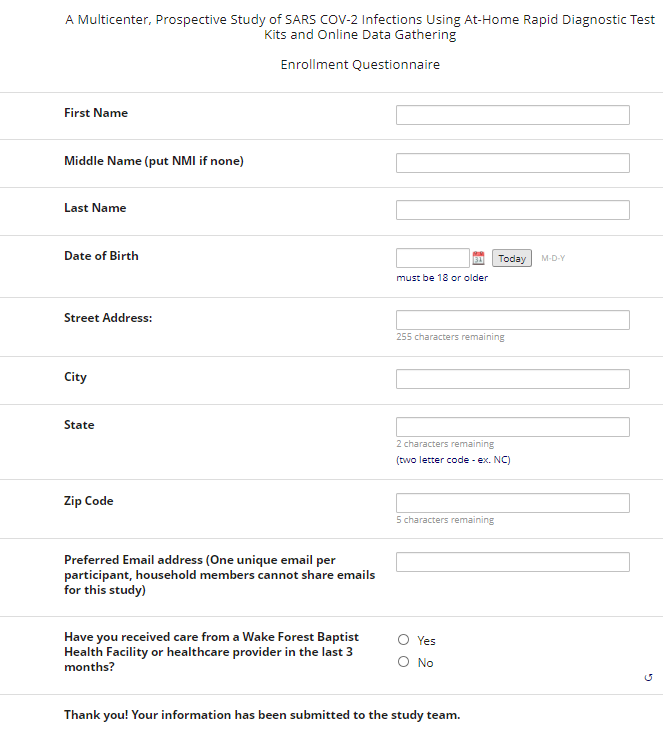

### Supplemental Figure 3

Supplemental Figure 3: Registration and Demographics Form

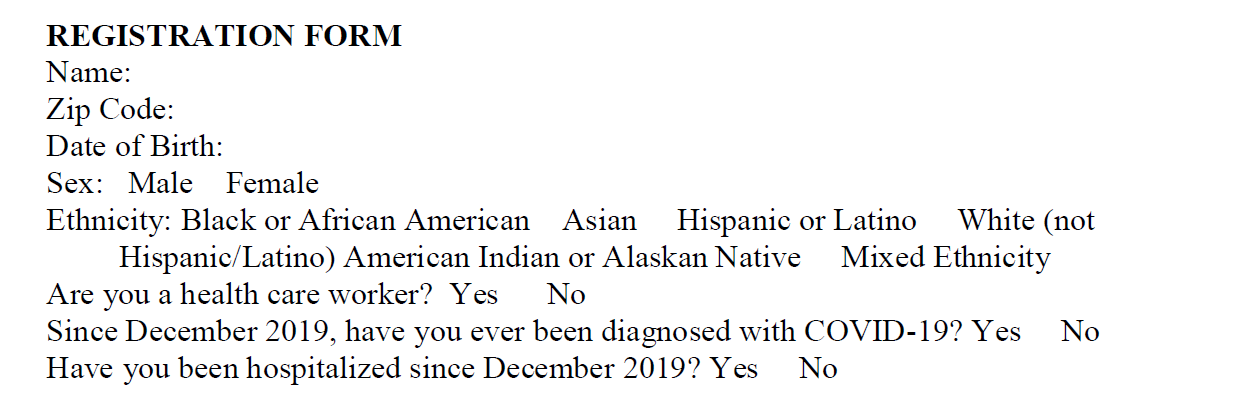

### Supplemental Figure 4

Supplemental Figure 4: Daily COVID-like Illness Report

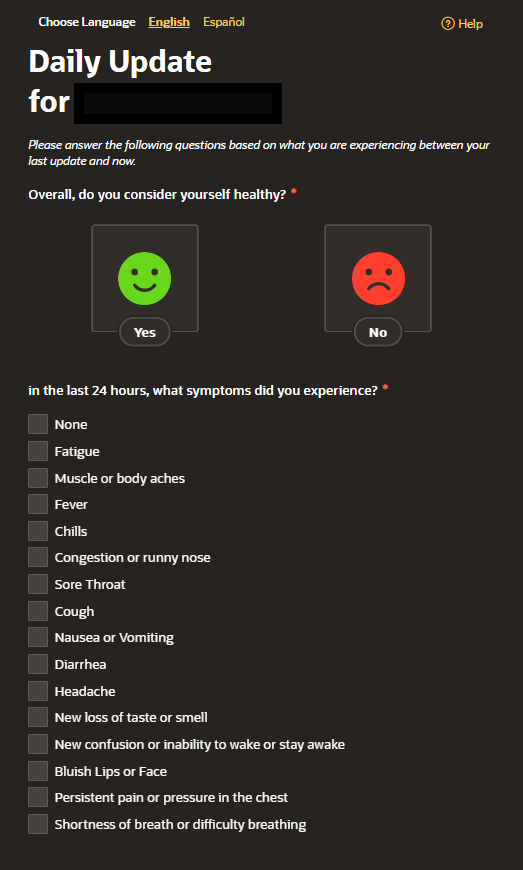

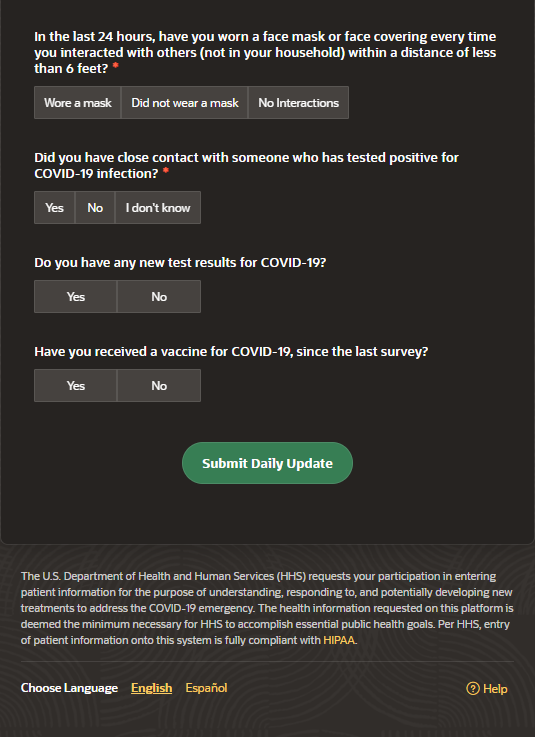
