## Supplemental Table 1 for "The COVID-19 Community Research Partnership: Objectives, Study Design, Baseline Recruitment, and Retention"

| Table 1: COVID-19 Community Research Partnership Collaborating Health Systems/Centers, Participant Eligibility Criteria and Enrollment Strategy. | | | | |
| --- | --- | --- | --- | --- |
|  | Health System or Center | Characteristics | Eligibility Criteria | Enrollment Strategy |
| 1. | Wake Forest Baptist Health | WFBH serves 3 million lives in the Piedmont Triad of northwest North Carolina including the cities of Greensboro, Winston-Salem, and High Point. The WFBH system includes Wake Forest Baptist Medical Center, an 855-bed tertiary care facility in Winston Salem that includes Brenner Children’s Hospital, five community hospitals, more than 350 primary and specialty care locations with more than 2,500 physicians and Wake Forest School of Medicine. | Began the study cohort of only health care workers and clients Wake Forest Baptist Health clinic locations. Health care workers who did not seek medical services through WFBH were not enrolled initially. As the study progressed, enrolment was opened to all adults (18 years or older) in October 2020. Participant decision to not provide informed consent was the only exclusion criteria.  A secondary pediatric cohort was also added to the study late in December 2020. Only children ages 2-17 were eligible for participation. | At the start of the study, only healthcare workers and clients of the Wake Forest Baptist Health system were eligible for enrollment in the CRP study. Messaging via study advertisements, email, and the electronic patient portal “MyWakeHealth” were all used for the first rounds of recruitment. MyWakeHealth varied to target healthcare workers separately from patients. Once enrollment was open to all adults, a generic public-facing website was established to show general study information and link potential participants to an enrollment questionnaire and informed consent if they wished to join the study. Community volunteers were also added to help recruit individuals from underserved and diverse communities around the WFBH-supported area.  For pediatric enrollment, messages were sent via MyWakeHealth to guardians enrolled in the CCRP asking if they wished to enroll their adolescent or young child in the study. |
| 2. | Atrium Health | Formerly known as Carolinas HealthCare System, Atrium Health is a not-for-profit hospital network with operates hospitals, freestanding emergency departments, urgent care centers and medical practices in North Carolina, South Carolina, and Georgia. | Adult: No exclusionary criteria other than unwillingness to give consent.  Pediatric: No exclusionary criteria other than unwillingness for parent to give consent. | Adult: Information about the study was emailed to all Atrium Health employees (~60k) and all Atrium Health patients (~900k). In addition, flyers were shared at clinics, mass vaccination events and in discharge instructions. Details of the study were also provided to local news outlets as well as through the MyAtriumHealth patient portal, and the Atrium Health website Healthbot. Approximately 17000 postcards were mailed to patients from Atrium Health clinics that serve large racial/ethnic minority populations based on geographic location. Flyers were distributed from Atrium mobile health units.  Atrium research team members worked with the Atrium Health Diversity and Inclusion department to facilitate community engagement. Social media and town hall events took place in early 2021 to discuss the study among other topics, including COVID-19 and vaccine hesitancy. COVID care packages, with flyers, masks and hand sanitizer, were distributed at community outreach events.  The recruitment team has also developed educational videos on COVID-19 which provide an overview of the study, myths surrounding COVID-19, benefits to participating in the study, and how the data will be used. The videos have been produced by Picnic Table Productions and are available in English and Spanish. The videos are finalized and are now live on the study website for participants to view.  Pediatric: Recruitment efforts for pediatric participants has been very similar to adults, with town hall events, distribution of flyers, and community outreach events. |
| 3. | Tulane University School of Public Health (SPH) and Tropical Medicine | Tulane recruited participants from three health networks that serve New Orleans and southeast Louisiana: Tulane University Medical Group (TUMG), Access Health Louisiana (AHL), and DePaul Community Health Centers (DCHC). | Individuals were eligible to enroll if they were a patient or employee of one of the three associated health networks and were 18 years of age or older. | Participants were enrolled through the Tulane CRP study website. The website has information about the study and a health network-specific link to the SneezSafe website where potential participants could complete the enrollment questionnaire and sign the informed consent. The major enrollment pushes occurred through rounds of email, text, and automated phone messages from the health networks to their eligible patient population with information about the study and a link to the Tulane CRP study website. Flyers and posters, digital and printed, were placed in clinics with a QR code and link for the Tulane CRP study website. Recruitment messages and materials were IRB-approved and given to the sites to utilize. |
| 4. | University of Mississippi Medical Center | UMMC provides wide-ranging patient care programs, including a children's hospital, a women and infants' hospital, and a critical care hospital, along with University Hospital. University Physicians includes about 500 doctors, many of them among leaders in their field, who care for patients in the hospitals and clinics on campus, around the Jackson metro area, and in outreach clinics around the state. UMMC includes two community hospitals, UMMC Holmes County and UMMC Grenada. | Patients and employees within the UMMC healthcare system aged 18 or older at time of enrollment. | Study advertisements, email, and messaging via electronic patient portal (MyChart) were utilized to recruit UMMC patients and employees. Interested participants reviewed study information and completed consent form online. |
| 5. | MedStar Health | MedStar Health is a healthcare leader in Maryland, Virginia, and the Washington, D.C. area with a partnership with George Washington University. MedStar is comprised of 30,000 associates and 5,400 affiliated physicians across 10 hospitals, numerous ambulatory care practices, and urgent care facilities. It is the largest provider of healthcare services in the region. | Eligible participants must meet all the following criteria: ability to provide informed consent, age 18 or older, have a valid email address, and able to communicate in English or Spanish. | MedStar transmitted 3 million study marketing correspondences. The primary method was MedStar Health’s Corporate email pool of unique individuals who at some point received health care and/or provided their personal email for communications with the system. The overall strategy included community press releases, social media campaigns, interviews, town halls, round table discussions with associates, grand rounds with providers, system-wide internal banners, messaging, flyers, bulletins, reminders, patient advisory boards and community groups. |
| 6. | University of Maryland | The University of Maryland Medical Center (UMMC) is the flagship academic medical center at the heart of University of Maryland Medical System (UMMS) and includes the 789-bed downtown Baltimore campus and the 177-bed midtown campus one mile north. The medical staff comprises nearly 1,200 attending physicians who are faculty members at the University of Maryland School of Medicine, as well as 900 residents and fellows in all medical specialties. | Primary eligibility included all UMMS patients aged 18 or older at the time of enrollment. Secondary study populations included community individuals who were not current patients of UMMS. Participants were also eligible to enroll without invitation through the study website. | Recruitment was initiated via emails to UMMS patients with an electronic health record (MyPortfolio). The message included a link to our study site, which contained a link to our REDCap consent and enrollment forms. Emails were sent in batches of approximately 9,000 to 21,000 to groups of patients at various age groups (oldest to youngest). Recruitment also included the use of emails, flyers, and social media campaigns. We used targeted marketing, interaction with community leaders, and interviews/webinars to reach special populations. We shared study information with the MD Department of Health which distributed information in some of their health clinics. We participated in a radio show with the University of Maryland College Park Center for Health Equity of College Park in an effort to have a wide reach in the African American community in the Washington D.C. Metropolitan area. We partnered with a local community and faith-based organization, Sisters Together and Reaching, to share information about the COVID-19 CRP via their social media platforms and throughout their networks. We also connected with a community-based organization that works with homeless youth. Our strategies with specific tactics to target young African Americans included outreach to local Historically Black Colleges and Universities as well as outreach to Black churches in the Baltimore area. To round out our efforts to recruit participants from the local community who were not affiliated with UMMS, we sent over 5000 recruitment emails to individuals who have expressed interested in research studies at the UMB Center for Vaccine Development.  We collaborated with MedStar to conduct a joint Town Hall for participants to provide study updates, general information about COVID-19 and information about COVID-19 vaccines in which over 400 participants joined. |
| 7. | New Hanover Regional Medical Center | New Hanover Regional Medical Center (NHRMC) is a not-for-profit 800-bed teaching hospital, regional referral center, and UNC School of Medicine branch campus. NHRMC serves 7 counties in Southeastern North Carolina and is nationally accredited by DNV GL – Healthcare. | Adults ≥ 18 years of age who receive health care at NHRMC were included. Focus groups included healthcare workers and patients diagnosed with cancer who received immune compromising cancer therapy during the pandemic. | A study webpage was developed and made accessible to members of the community. Information about the study was presented to organizational leaders during our monthly leadership meeting. Announcements were provided in multiple streams of communication: the internal employee communication website, the daily email to internal employees, and the ‘News & Notes’ email to internal medical staff, A press release was delivered and interviews given with local news reporters. |
| 8. | Vidant Health | Vidant Health is a mission-driven, 1,447-bed **health** system serving more than 1.4 million people in 29 eastern North Carolina counties. The not-for-profit system is made up of 12,000 employees, eight hospitals, home **health, hospital, wellness centers and Vidant Medical Group.** | Adult patients 18 years and older were included. We focused primarily on those who had been patients of the Vidant Health system, but were willing to sign up anyone who wanted to participate. | Multiple methods of communication were employed, including email, EHR messaging, digital ads on the Vidant Health webpage, and community events (including local churches). We also hired 20 community health advocates to contact locations (stores, voting stations, hospital lobby) and set up booths to attract new participants. |
| 9. | WakeMed Health & Hospitals | WakeMed is a 941-bed private, not-for-profit health care system based in Raleigh, North Carolina. | English-speaking adults age 18 years and older were included. English-speaking children age 2-17 were also included. Participants were not required to be a patient of WakeMed. | Adults: A recruitment message from the chief medical officer (CMO) was sent to all English-speaking WakeMed patients with an active MyChart account. Similarly, a recruitment email from the CMO was sent to all WakeMed staff and providers. A series of advertisements appeared in WakeMed Weekly, a weekly email newsletter to all staff and providers. The study was advertised in Microscope, a monthly printed employee newsletter. Social media platforms (Instagram, Facebook) were used to post information about the study. A fixed advertisement appeared on the WakeMed intranet homepage.  Children: An email was sent to all enrolled adults to recruit children in their households. A series of social media (Instagram, Facebook) advertisements were posted. Flyers were available in WakeMed Pediatric Primary Care and pediatric sub-specialties clinics. An email with study information was sent to all WakeMed pediatric community partners and providers.  Advertisements appeared in WakeMed Weekly and ‘A Word from WakeMed Childrens’, a newsletter from WakeMed Childrens’ to community pediatric providers. Lastly, an informational slide was included in a rolling PowerPoint presentation at a local COVID-19 vaccine clinic observation room. |
| 10. | Campbell University School of Osteopathic Medicine | The School is located in rural North Carolina and works with additional sites including but not limited to Harnett Health, Cape Fear Valley Medical Center, Southeastern Regional Medical Center, and Sampson Regional Medical Center to enroll underserved and underrepresented populations. | The study was open to the entire population. No restrictions were in place for participation. | Sites working with Campbell University utilized the health system’s electronic messaging portal “Mychart Messaging”. Many sites also distributed paper flyers, published study information in a local health newsletter, and conducted in-person recruiting activities. |
