## Supplemental Table 2 for "The COVID-19 Community Research Partnership: Objectives, Study Design, Baseline Recruitment, and Retention"

**Electronic Health Records (EHR)**

**Data to Extract from EHR Systems**

The following document lists data to be extracted from each participating sites EHR. The data elements are organized into four categories: protected health information (PHI) needed for data merging and serologic sampling; domain 1: demographics and behavioral information; domain 2: hospitalizations, ambulatory encounters, diagnoses, and measurements; and domain 3: all active medications. Each data element is defined in the EHR data dictionary which further describes the elements including the source, format, and associated codes for common data models. The EHR data dictionary is currently in progress and will be updated iteratively as the data extraction process progresses.

Data will be transferred from each site to the Biostatistics Center’s Server using secure FTP. The active problem list and active medication list will be extracted, along with past problems and medications, to allow analysis of the specific data listed in domains 2 and 3. Therefore, the data elements listed in domains 2 and 3 are sample elements that highlight areas of interest and not meant to represent an exhaustive list.

Concerning active medications, all data is pulled. Active medications are defined as any medication taken during the study period. We will delineate between patient administered medications vs facility administered medications. Additionally, we need to include all transfusions including convalescent plasma, etc.

**Protected Health Information (PHI) Needed for Data Merge (and Serologic Sampling)**

1. Name
2. Address including zip code
3. Date of Birth
4. Email addresses (x2 if available)
5. Phone number
6. Unique system identifier i.e. medical record number (we need to identify a way to ensure HCS with multiple facilities can distinguish between multiple sites to avoid MRN collisions)
7. Internal Study Identifier

**Domain 1: Demographics and behavioral information**

1. Race/ethnicity
2. Gender
3. Height and Weight
4. Type of insurance
5. Employment status
6. Smoking status
7. Alcohol use/drug use via coded problems

**Domain 2: Hospitalizations, Ambulatory Encounters, Diagnoses, Measurements.**

1. Hospitalizations and Ambulatory Encounters
   a. Diagnoses
   b. Dates, Length of stay
   c. Outcomes (discharged vs deceased)
   d. Discharge disposition
   e. Cause of death (may require manual process after identifying death)
2. Proxies for Intensive Care Unit (ICU) usage or level of care (specific to hospitalizations)
   a. Ventilation
   b. Extracorporeal membrane oxygenation (ECMO)
   c. Level of oxygen support (intubation, cannulation, high-flow nasal cannula (HFNC), oxygen flow rate)
   d. Bed status orders
3. Vitals
   a. Blood pressure
   b. Heart rate
   c. Respiratory rate
   d. Temperature
4. Oxygen saturation (i.e. pulse ox)
5. Active problem list
   a. Hypertension
   b. Diabetes
   c. Coronary artery disease, stroke or heart failure
   d. Chronic liver disease (cirrhosis)
   e. Hypercoagulability, clotting issues
   f. Lung disease
      i. Asthma
      ii. Chronic obstructive pulmonary disease (COPD)
   g. Obstructive sleep apnea (OSA)
   h. Cancer
      i. Solid tumor
      ii. Leukemia/lymphoma
   i. Chronic kidney disease + stage, if known
   j. Human immunodeficiency virus (HIV)
   k. Transplant recipient
      i. Solid organ
      ii. Stem cell/bone marrow
   l. Autoimmune disorder (e.g. lupus, Crohn’s, rheumatoid arthritis)
   m. Multisystem Inflammatory Syndrome Children (MISC)
   n. Multisystem Inflammatory Syndrome Adults (MISA)
   o. Pregnancy status
   p. Mental health diagnoses
6. Laboratory testing results (including order and result date, and assay type/manufacturer)
   a. COVID-19
      i. Reverse transcription-polymerase chain reaction (RT-PCR)
      ii. Antigen
      iii. Antibody (Spike or Nucleocapsid; Immunoglobulins (IgA, IgA, IgM, or a combination))
   b. HIV test results and laboratory markers
      i. Viral load (VL)
      ii. Cluster of differentiation 4 (CD4)
      iii. Chem screen (CS)
      iv. Complete blood count (CBC)
   c. Inflammatory markers (erythrocyte sedimentation rate (ESR), C-reactive protein (CRP) ferritin, lactase dehydrogenase (LDH))
   d. Coagulation markers (prothrombin time (PT), partial thromboplastin time (PTT), international normalized ratio (INR), D-dimer, fibrinogen)
   e. Kidney function tests (blood urea nitrogen (BUN), creatinine (Cr))
   f. Liver function tests (aspartate transaminase (AST), alanine transaminase (ALT), alkaline phosphatase (ALP), gamma-glutamyl transferase (GGT), Albumin)
7. Immunization history including vaccine type, manufacturer, and dose

**Domain 3: All Active Medications**

1. Cardiovascular System – Agents Acting on the Renin-Angiotensin System:
   a. Angiotensin converting enzyme (ACE) inhibitor
   b. Angiotensin receptor blocker (ARB)
2. Cardiovascular System – Lipid modifying agents:
   a. HMG-CoA Reductase inhibitor (statin)
3. Cardiovascular System – Diuretics:
   a. Congestive Heart Failure (i.e. Lasix and Bimex)
4. Cardiovascular System – antihypertensives:
5. Alimentary Tract and Metabolism – Drugs used in Diabetes:
   a. Diabetes medications (such as insulin)
   b. Dipeptidyl peptidase 4 (DPP-4) inhibitors
   c. Dapagliflozin
6. Alimentary Tract and Metabolism – Drugs for acid related disorders:
   a. Proton pump inhibitor (PPI)
   b. H2-Antagonists
   c. Famotidine
7. Antineoplastic and Immunomodulating Agents – antineoplastic:
   a. Methotrexate
8. Antineoplastic and Immunomodulating Agents - immunosuppressants :
   a. Mycophenolate
   b. Cyclosporine
   c. Tacrolimus
   d. Monoclonal antibody infusions (e.g. TNF-alpha inhibitors, tocilizumab, adalimumab, anakinra)
   e. IL-23 inhibitors (risankizumab, guselkumab, and tildrakizumab)
   f. Interferon beta-1a
9. Antineoplastic and Immunomodulating Agents - selective immunosuppressants
   a. Janus kinase (JAK) inhibitors (e.g. tofacitinib, baricitinib)
10. Ophthalmological and ontological preparations – anti-infectives and corticosteroids:
    a. Corticosteroid
11. Respiratory system – Nasal Preparations:
    a. Inhaled corticosteroids
12. Blood and Blood Forming Agents – Antithrombotics:
    a. Warfarin
    b. Direct oral anticoagulants (DOACs)
    c. Low-molecular weight heparin
    d. Unfractionated heparin
    e. Aspirin
13. Anti-Infectives for Systemic Use – Antivirals:
    a. Lopinavir
    b. Ritonavir
    c. Darunavir
    d. Cobicistat
    e. Oseltamivir
14. Anti-Infectives for Systemic Use – Antibacterials:
    a. Azithromycin
15. Nervous System – Analgesics:
    a. Acetaminophen (paracetamol)
16. Antiparasitic products, insecticides, and repellents – antimalarials:
    a. Hydroxycloroquine

17. COVID-19 medications codes/other:

a. Hydroxychloroquine
b. Convalescent plasma
c. Remdesivir
