## Supplemental Table 3 for "The COVID-19 Community Research Partnership: Objectives, Study Design, Baseline Recruitment, and Retention"

Supplemental Table 2: Characteristics of Serologic Tests Utilized in the COVID-19 Community Research Partnership

| Assay | Target Antigen | Antibody | Performance Measure | Estimate of Performance | 95% Confidence Interval |
| --- | --- | --- | --- | --- | --- |
| EUROIMMUN SARS-COV-2 ELISA (IgG) | Spike | IgG | Sensitivity | 90.0% (27/30) | (74.4%; 96.5%) |
|  |  | IgG | Specificity | 100% (80/80) | (95.4%; 100%) |
|  |  | IgG | PPV at prevalence = 5% | 100% | (46.1%; 100%) |
|  |  | IgG | NPV at prevalence = 5% | 99.5% | (98.6%; 99.8%) |
| Roche Elecsys Anti-SARS-CoV-2 ECLIA | Nucleocapsid | Pan-Ig | Sensitivity (PPA) | 100% (29/29) | (88.3%; 100%) |
|  |  | Pan-Ig | Specificity (NPA) | 99.8% (5262/5272) | (99.7%; 99.9%) |
|  |  | Pan-Ig | PPV at prevalence = 5% | 96.5% | (93.0%; 98.1%) |
|  |  | Pan-Ig | NPV at prevalence = 5% | 100% | (99.4%; 100%) |
|  |  | Pan-Ig | Sensitivity (PPA) | 100% (29/29) | (88.3%; 100%) |
| Innovita 2019-nCoV Ab Test (Colloidal Gold) LFA | Spike and Nucleocapsid | IgM | Sensitivity | 80.0% (24/30) | (62.7%; 90.5%) |
|  |  | IgM | Specificity | 98.8% (79/80) | (93.3%; 99.8%) |
|  |  | IgG | Sensitivity | 83.3% (25/30) | (66.4%; 92.7%) |
|  |  | IgG | Specificity | 98.8% (79/80) | (93.3%; 99.8%) |
|  |  | IgM or IgG | Sensitivity | 90.0% (27/30) | (74.4%; 96.5%) |
|  |  | IgM or IgG | Specificity | 97.5% (78/80) | (91.3%; 99.3%) |
|  |  | IgM or IgG | PPV at prevalence = 5% | 65.5% | (31.1%; 88.1%) |
|  |  | IgM or IgG | NPV at prevalence = 5% | 99.5% | (98.5%; 99.8%) |

**ELISA =** enzyme-linked immunosorbent assay

**ECLIA =** electro-chemiluminescence immunoassays

**LFA** = lateral flow assay
