## Supplemental Table 4 for "The COVID-19 Community Research Partnership: Objectives, Study Design, Baseline Recruitment, and Retention"

Supplemental Table 3: Demographics by Study Sites

A) National Coalition

|  | **Number (Percent) Enrolled in NC State Wake Forest** | **Number (Percent) Enrolled in NC State Atrium** | **Number (Percent) Enrolled in NC State Campbell** | **Number (Percent) Enrolled in NC State New Hanover** | **Number (Percent) Enrolled in NC State Vidant Health** | **Number (Percent) Enrolled in NC State Wake Med** | **Number (Percent) Enrolled in NC State Total** |
| --- | --- | --- | --- | --- | --- | --- | --- |
| **Total** | 12220 | 4099 | 536 | 761 | 990 | 3243 | 21849 |
| **Age** |  |  |  |  |  |  |  |
| 18-29 | 1016 (8.3%) | 310 (7.6%) | 77 (14.4%) | 66 (8.7%) | 60 (6.1%) | 343 (10.6%) | 1872 (8.6%) |
| 30-39 | 2307 (18.9%) | 818 (20.0%) | 101 (18.8%) | 148 (19.4%) | 118 (11.9%) | 566 (17.5%) | 4058 (18.6%) |
| 40-49 | 2508 (20.5%) | 1018 (24.8%) | 128 (23.9%) | 165 (21.7%) | 158 (16.0%) | 819 (25.3%) | 4796 (22.0%) |
| 50-59 | 2734 (22.4%) | 1106 (27.0%) | 125 (23.3%) | 149 (19.6%) | 237 (23.9%) | 640 (19.7%) | 4991 (22.8%) |
| 60-69 | 2472 (20.2%) | 659 (16.1%) | 76 (14.2%) | 154 (20.2%) | 242 (24.4%) | 853 (26.3%) | 4456 (20.4%) |
| 70-79 | 1044 (8.5%) | 160 (3.9%) | 29 (5.4%) | 73 (9.6%) | 149 (15.1%) | 21 (0.6%) | 1476 (6.8%) |
| >=80 | 139 (1.1%) | 28 (0.7%) | 0 (0.0%) | 6 (0.8%) | 26 (2.6%) | 1 (0.0%) | 200 (0.9%) |
| **Sex** |  |  |  |  |  |  |  |
| Female | 8679 (71.0%) | 3357 (81.9%) | 421 (78.5%) | 574 (75.4%) | 722 (72.9%) | 2221 (68.5%) | 15974 (73.1%) |
| Male | 3533 (28.9%) | 742 (18.1%) | 115 (21.5%) | 177 (23.3%) | 268 (27.1%) | 893 (27.5%) | 5728 (26.2%) |
| Not Declared | 8 (0.1%) | 0 (0.0%) | 0 (0.0%) | 10 (1.3%) | 0 (0.0%) | 129 (4.0%) | 147 (0.7%) |
| **Race/ethnicity** |  |  |  |  |  |  |  |
| White (not Hispanic/Latino) | 11886 (97.3%) | 3925 (95.8%) | 386 (72.0%) | 676 (88.8%) | 822 (83.0%) | 2651 (81.7%) | 20346 (93.1%) |
| Black or African American | 144 (1.2%) | 62 (1.5%) | 50 (9.3%) | 19 (2.5%) | 102 (10.3%) | 173 (5.3%) | 550 (2.5%) |
| American Indian or Alaskan | 8 (0.1%) | 3 (0.1%) | 58 (10.8%) | 1 (0.1%) | 7 (0.7%) | 9 (0.3%) | 86 (0.4%) |
| Asian or Pacific Islander | 49 (0.4%) | 27 (0.7%) | 11 (2.1%) | 10 (1.3%) | 11 (1.1%) | 86 (2.7%) | 194 (0.9%) |
| Hispanic or Latino | 62 (0.5%) | 33 (0.8%) | 15 (2.8%) | 22 (2.9%) | 17 (1.7%) | 82 (2.5%) | 231 (1.1%) |
| Mixed Ethnicity | 33 (0.3%) | 16 (0.4%) | 6 (1.1%) | 14 (1.8%) | 21 (2.1%) | 73 (2.3%) | 163 (0.7%) |
| Not Declared/Specified | 38 (0.3%) | 33 (0.8%) | 10 (1.9%) | 19 (2.5%) | 10 (1.0%) | 169 (5.2%) | 279 (1.3%) |

B) North Carolina State Coalition Sites

|  | **Number (Percent) Enrolled in NC State Wake Forest** | **Number (Percent) Enrolled in NC State Atrium** | **Number (Percent) Enrolled in NC State Campbell** | **Number (Percent) Enrolled in NC State New Hanover** | **Number (Percent) Enrolled in NC State Vidant Health** | **Number (Percent) Enrolled in NC State Wake Med** | **Number (Percent) Enrolled in NC State Total** |
| --- | --- | --- | --- | --- | --- | --- | --- |
| **Total** | 12220 | 4099 | 536 | 761 | 990 | 3243 | 21849 |
| Age |  |  |  |  |  |  |  |
| 18-29 | 1016 (8.3%) | 310 (7.6%) | 77 (14.4%) | 66 (8.7%) | 60 (6.1%) | 343 (10.6%) | 1872 (8.6%) |
| 30-39 | 2307 (18.9%) | 818 (20.0%) | 101 (18.8%) | 148 (19.4%) | 118 (11.9%) | 566 (17.5%) | 4058 (18.6%) |
| 40-49 | 2508 (20.5%) | 1018 (24.8%) | 128 (23.9%) | 165 (21.7%) | 158 (16.0%) | 819 (25.3%) | 4796 (22.0%) |
| 50-59 | 2734 (22.4%) | 1106 (27.0%) | 125 (23.3%) | 149 (19.6%) | 237 (23.9%) | 640 (19.7%) | 4991 (22.8%) |
| 60-69 | 2472 (20.2%) | 659 (16.1%) | 76 (14.2%) | 154 (20.2%) | 242 (24.4%) | 853 (26.3%) | 4456 (20.4%) |
| 70-79 | 1044 (8.5%) | 160 (3.9%) | 29 (5.4%) | 73 (9.6%) | 149 (15.1%) | 21 (0.6%) | 1476 (6.8%) |
| >=80 | 139 (1.1%) | 28 (0.7%) | 0 (0.0%) | 6 (0.8%) | 26 (2.6%) | 1 (0.0%) | 200 (0.9%) |
| **Sex** |  |  |  |  |  |  |  |
| Female | 8679 (71.0%) | 3357 (81.9%) | 421 (78.5%) | 574 (75.4%) | 722 (72.9%) | 2221 (68.5%) | 15974 (73.1%) |
| Male | 3533 (28.9%) | 742 (18.1%) | 115 (21.5%) | 177 (23.3%) | 268 (27.1%) | 893 (27.5%) | 5728 (26.2%) |
| Not Declared | 8 (0.1%) | 0 (0.0%) | 0 (0.0%) | 10 (1.3%) | 0 (0.0%) | 129 (4.0%) | 147 (0.7%) |
| **Race/ethnicity** |  |  |  |  |  |  |  |
| White (not Hispanic/Latino) | 11886 (97.3%) | 3925 (95.8%) | 386 (72.0%) | 676 (88.8%) | 822 (83.0%) | 2651 (81.7%) | 20346 (93.1%) |
| Black or African American | 144 (1.2%) | 62 (1.5%) | 50 (9.3%) | 19 (2.5%) | 102 (10.3%) | 173 (5.3%) | 550 (2.5%) |
| American Indian or Alaskan | 8 (0.1%) | 3 (0.1%) | 58 (10.8%) | 1 (0.1%) | 7 (0.7%) | 9 (0.3%) | 86 (0.4%) |
| Asian or Pacific Islander | 49 (0.4%) | 27 (0.7%) | 11 (2.1%) | 10 (1.3%) | 11 (1.1%) | 86 (2.7%) | 194 (0.9%) |
| Hispanic or Latino | 62 (0.5%) | 33 (0.8%) | 15 (2.8%) | 22 (2.9%) | 17 (1.7%) | 82 (2.5%) | 231 (1.1%) |
| Mixed Ethnicity | 33 (0.3%) | 16 (0.4%) | 6 (1.1%) | 14 (1.8%) | 21 (2.1%) | 73 (2.3%) | 163 (0.7%) |
| Not Declared/Specified | 38 (0.3%) | 33 (0.8%) | 10 (1.9%) | 19 (2.5%) | 10 (1.0%) | 169 (5.2%) | 279 (1.3%) |
